# The impact of attending rheumatologist’s Big Five personality traits on systemic lupus erythematosus patients’ trust in their rheumatologist: the TRUMP^2^-SLE project

**DOI:** 10.1101/2023.05.22.23290279

**Authors:** Nao Oguro, Nobuyuki Yajima, Yuichi Ishikawa, Natsuki Sakurai, Chiharu Hidekawa, Takanori Ichikawa, Dai Kishida, Keigo Hayashi, Kenta Shidahara, Yoshia Miyawaki, Ryusuke Yoshimi, Ken-ei Sada, Yasuhiro Shimojima, Noriaki Kurita

## Abstract

**Objectives:** Differences in communication styles according to physicians’ personality traits have been identified, and such physician-related factors can be important in building patient-physician trust. This study examined the impact of rheumatologists’ Big Five personality traits on patients’ trust in their attending rheumatologists.

**Methods:** This cross-sectional study included Japanese patients with systemic lupus erythematosus (SLE) at five academic medical centres between June 2020 and August 2021. The exposure was the Big Five personality traits of their attending rheumatologists using the Japanese version of the Ten-Item Personality Inventory Scale (2-14 points each). The outcome was the patient’s trust in attending rheumatologist using the Japanese version of the 5-item Wake Forest Physician Trust Scale (0-100 points). A general linear model was fitted to adjust for rheumatologists’ and patients’ characteristics, while correcting for the clustering effect of the same attending rheumatologists.

**Results:** The study included 505 patients with a mean age of 46.8 years, and 88.1% were women. Forty-three attending rheumatologists (mean age, 39.6 years, 23.3% women) were identified. Higher extraversion and agreeableness were associated with higher trust in attending rheumatologists (per 1-point increase, 1.85 points [95% CI 0.52 –3.19] and 2.24 points [95% CI 0.86 –3.61], respectively), and higher conscientiousness was associated with lower trust (per 1-point increase, −1.07 points [95% CI −1.64 – −0.49]).

**Conclusions:** While higher extraversion and agreeableness of attending rheumatologists led to higher patient trust in their rheumatologist, overly high conscientiousness may lead to lower trust resulting from the physician’s demand of dutifulness from patients with SLE.

**KEY MESSAGES:** **What is already known on this topic**

Although physicians’ personality traits are known to influence their attitudes and performance, little is known about how they affect trust through rheumatology patient-physician interactions.

**What this study adds**

Some Big Five personality traits of an attending rheumatologist were associated with patients’ trust in their rheumatologist; extraversion and agreeableness were linked to higher trust and conscientiousness was associated with lower trust.

**How this study may affect research, practice, or policy**

Through reflection on their personality traits, attending rheumatologists are expected to become aware of their habits in patient-physician interactions and adapt their communication to strengthen the relationships with their patients.

## INTRODUCTION

The patient-physician trusted relationship constitutes the cornerstone of patient-centred care in rheumatology practice [1] and is an important source of empowerment for patients with systemic lupus erythematosus (SLE). [2,3] Strong trusted relationships can lead to improvements in clinical outcomes,[4] and higher levels of SLE patients’ trust in their attending physician are associated with better adherence to medications. [3,5] Therefore, methods for enhancing trust in physicians are important in the SLE management. To date, studies on SLE have shown that health literacy [6] and psychological factors, such as hope [3] on the patient’s part and misdiagnosis on the physician’s part, have an impact on patients’ trust in their physicians. [2,7,8] However, there is insufficient evidence regarding the influence of attending physicians’ personality traits, which affects their communication styles, on the formation of trust.

Physicians’ personality characteristics have been focused mainly on medical education [9,10]; in particular, the Big Five personality traits (also known as the five-factor model of personality) consisting of extraversion, agreeableness, openness, conscientiousness, and emotional stability have been examined. [11–13] A study examining physician personality characteristics associated with physician satisfaction ratings showed that a high level of openness, linked to imagination and intellectual curiosity, [13] was associated with greater patient satisfaction, suggesting the importance of an empathetic response and an atmosphere that easily allows for discussion. [14] Moreover, an average level, rather than overly high levels, of physician conscientiousness was associated with higher physician ratings. However, how the attending physician’s personality affects “trustworthiness”, reflecting expectations for the future as opposed to previous physician ratings, remains unclear. Studying the impact of specific physician personality traits may be useful in devising strategies to improve the care process. This significance is supported by interviews of SLE patients who have highlighted the importance of being a “personable” physician in physician-patient communication and trusted relationship. [2,15]

Therefore, the present study, using cross-sectional data from the multicentre; the Trust Measurement for Physicians and Patients with SLE (TRUMP^2^-SLE) project, examined how the Big Five personality traits of attending physicians impact physician trust among patients with SLE.

## METHODS

### Study design and setting

This cross-sectional analysis used baseline information from the TRUMP^2^-SLE project, an ongoing multicentre cohort study conducted at five academic medical sites (Showa University Hospital, Okayama University Hospital, Shinshu University Hospital, Yokohama City University Hospital, and Yokohama City University Medical Center). This study was performed in accordance with the Declaration of Helsinki and Good Clinical Practice guidelines, and was approved by the Ethics Review Board of Showa University (22-298-B).

The inclusion criteria were as follows: (1) SLE diagnosis, according to the revised 1997 American College of Rheumatology classification criteria, (2) aged 20 years or older, (3) receiving rheumatology care at the participating centre, and (4) able to respond to the questionnaire survey. Patients with dementia or total blindness were excluded. All rheumatologists who treated the patients were Japanese. Rheumatologists participating in the TRUMP^2^-SLE trial were recruited from the participating hospitals and they voluntarily completed self-reported questionnaires. The attending rheumatologists of patients enrolled in the TRUMP^2^-SLE trial were included, regardless of whether they were research investigators. The response rate was set at 100%.

### Exposure

Physicians’ personality traits were measured using the Japanese version of the Ten-Item Personality Inventory (TIPI-J) scale, [16,17] which measures the Big 5 personality traits, a generally accepted taxonomy of personality traits (Supplementary Table 1). The TIPI-J scale consists of 10 items, with two questions for each domain, and is scored on a seven-point Likert scale. The five domains comprised extraversion, agreeableness, conscientiousness, emotional stability, and openness. The Big Five personality dimensions–extroversion, agreeableness, openness, conscientiousness, and emotional stability–have been widely applied in research on personality traits. A high level of extroversion indicates sociability and emotional expressiveness, whereas a low level indicates introversion and shyness. A high level of agreeableness indicates kindness and altruism, whereas a low level indicates quarrelsome and prosocial behaviour. A high level of conscientiousness indicated orderliness and dutifulness, whereas a low level indicates careless and disorderly behaviour. A high level of emotional stability indicates stability and calmness, whereas a low level indicates anxiety and moodiness. A high level of openness indicates wide areas of interest and imagination, whereas a low level indicates commonplace and narrow interests.[18]

Doctors were instructed to score each item on a scale of 1 as “Disagree strongly” to 7 as “Agree strongly”, respectively. After reversing the scores for negatively worded items, each domain’s score was calculated by summing the item scores and ranged from two to 14 points. The TIPI-J scale was validated and demonstrated to have good reliability (alpha coefficients of 0.92, 0.85, 0.82, 0.91 and 0.86 for extraversion, agreeableness, conscientiousness, emotional stability, and openness, respectively) and construct validity.

### Outcome

The main outcome was “trust in one’s physician” which was measured by the Japanese version of the five-item Wake Forest Physician Trust Scale.[19,20] It comprised five items scored on a five-point Likert scale. Patients were asked to choose one response for each item ranging from “strongly disagree” (1 point) to “strongly agree” (5 points). After inverting the score for negatively worded items, the sum of the scores was converted into a scale of 0–100. To inquire about trust in their rheumatologist, the following instructive statement was presented: “Please answer these questions keeping in mind the physician who has recently treated you regularly with SLE. We refer to this physician as ‘your doctor’. For the next question, we were interested in the honest opinion of your doctor. For each of these questions, please state whether you strongly agree, agree, are neutral, disagree, or strongly disagree.” The coefficient alpha value for the Japanese version of the Interpersonal Trust in Physician Scale was 0.85, demonstrating construct validity.

### Measurement of covariates

The covariates included were patient age, sex, disease activity, duration of illness, doctor’s age [21], sex [22,23], employment position [23], and duration of the doctor’s treatment [7]. These covariates were chosen because they could be associated with trust in physicians and/or the physicians’ personality traits. Disease activity was measured by the attending physician using the Systemic Lupus Erythematosus Disease Activity Index 2000 (SLEDAI-2K). As there are only full professor and associate professor job titles in Japan that contain the Kanji character ‘kyouju’, meaning professor in English, and health information from an authority is more likely to be trusted, [24] the job titles were divided into associate professor or higher and lower. Questionnaires were administered to each facility between June 2020 and August 2022. Patients were allowed to complete the questionnaire either the waiting room or at home. The questionnaire included assurances that the attending physician would not view the responses, and that the responses would only be used at the central facility for aggregation.

### Statistical analysis

All statistical analyses were performed using Stata/SE, version 16.1 (StataCorp, College Station, TX, USA). Patient and rheumatologist characteristics were described as frequencies and proportions for categorical variables and means and standard deviations (SD) for continuous variables. Rheumatologists’ Big Five personality trait scores were compared with the representative values reported in the literature for the development of the TIPI-J, [16] with unpaired t-tests. A general linear model was fitted to examine the association between the’ Big Five personality traits of rheumatologists and patients’ trust in their physicians. In the model, explanatory variables included the five personality domains, physician characteristics (age, sex, and job title), patient characteristics (age, sex, disease duration, and SLEDAI-2K), and duration of the physician-patient relationship. To address the clustering of outcomes resulting from multiple patient ratings with similar trust levels for the same physician, a cluster-robust variance estimation was used, with each physician as the cluster unit. Missing data on covariates were addressed by multiple imputation. Twenty imputations were done using chained equations, with the assumption that the data were missing at random. To calculate the corresponding standardised ES (Cohen’s *d*), the point estimate was divided by the SD of the trust in the rheumatologist score. Statistical significance was set at p < 0.05.

### Patient and public involvement

The general public and patients with SLE did not participate in the design, recruitment, or execution of the study.

## RESULTS

### Study flow

In total, 521 SLE patients met the inclusion criteria. Of these, four did not provide their physicians’ names. After excluding 12 patients with missing data on outcomes, 505 patients were included in the analysis. Forty-three attending physicians participated in the study.

### Patient and physician characteristics

Patient characteristics are presented in Table 1. The mean age was 46.8 years (SD, 14.1) and 445 (88.1%) were women. Mean disease activity, as determined by the SLEDAI-2K scale, was 4.0 (SD 3.9) points. A total of 301 (62.8%) patients were treated by their attending rheumatologists for more than 3 years. The mean score of trust in one’s physician was 80.1 (SD, 16.0).

**Table 1.**
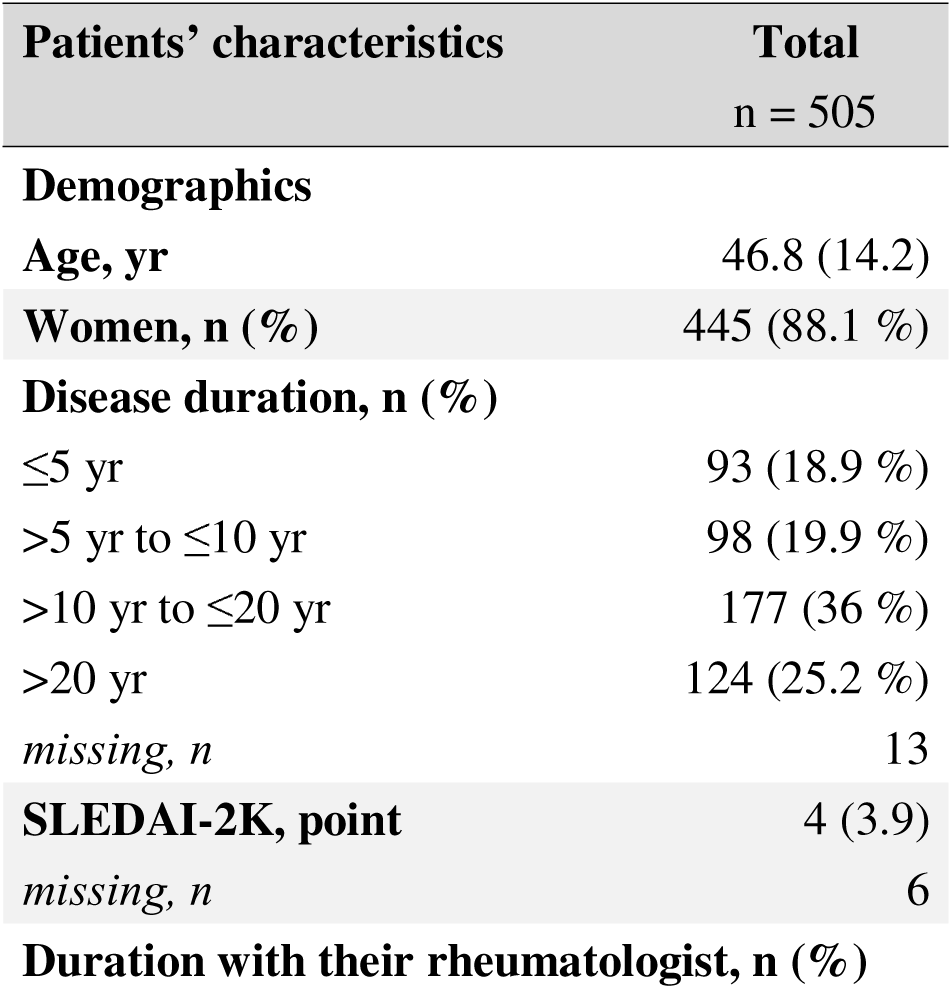
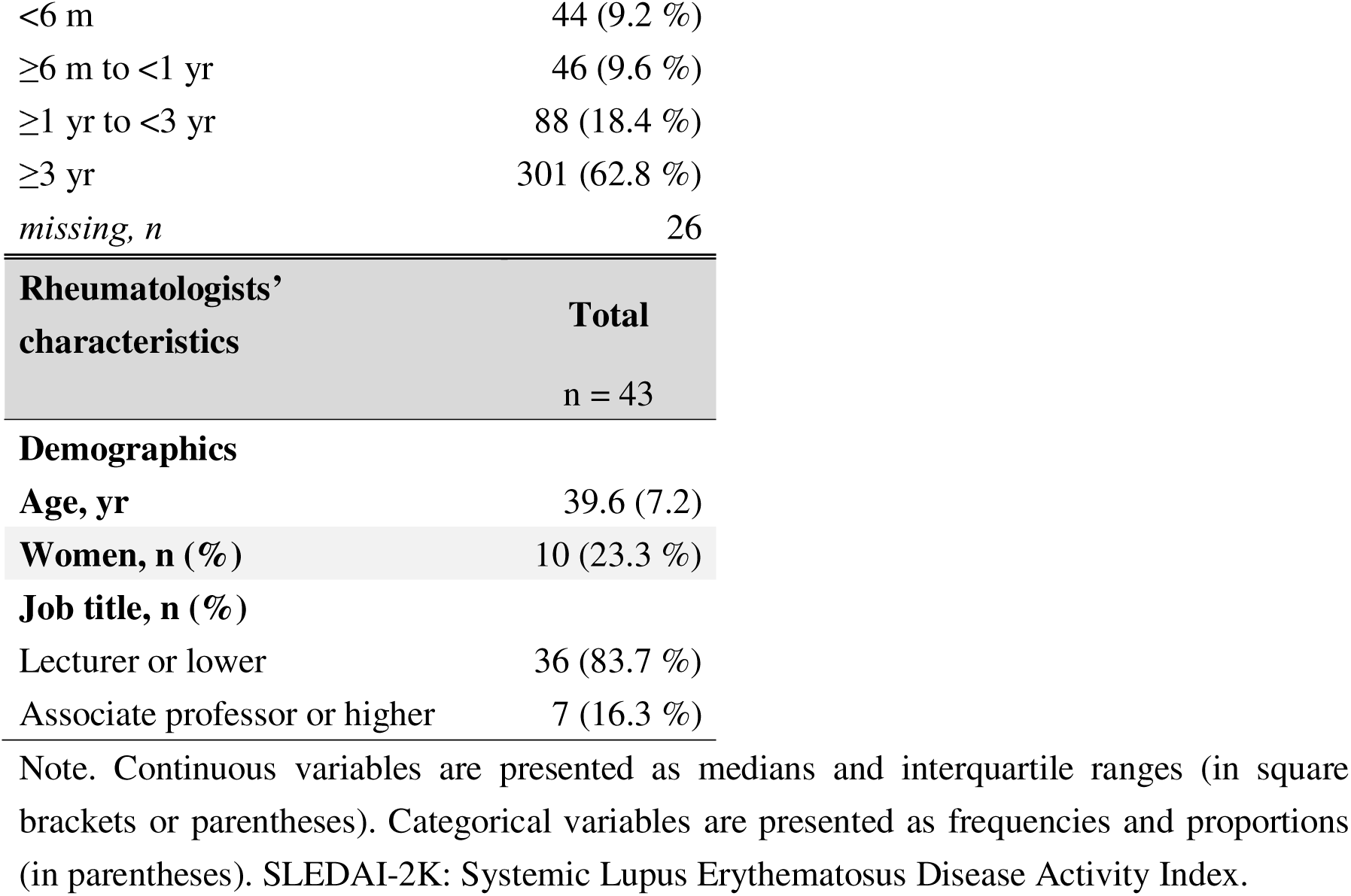
Patients’ and rheumatologists’ characteristics.

| **Patients' characteristics** | **Total**<br>**n = 505** |
| --- | --- |
| **Demographics** |  |
| **Age, yr** | 46.8 (14.2) |
| **Women, n (%)** | 445 (88.1 %) |
| **Disease duration, n (%)** |  |
| ≤5 yr | 93 (18.9 %) |
| >5 yr to ≤10 yr | 98 (19.9 %) |
| >10 yr to ≤20 yr | 177 (36 %) |
| >20 yr | 124 (25.2 %) |
| *missing, n* | 13 |
| **SLEDAI-2K, point** | 4 (3.9) |
| *missing, n* | 6 |
| **Duration with their rheumatologist, n (%)** |  |
| <6 m | 44 (9.2 %) |
| ≥6 m to <1 yr | 46 (9.6 %) |
| ≥1 yr to <3 yr | 88 (18.4 %) |
| ≥3 yr | 301 (62.8 %) |
| *missing, n* | 26 |

| **Rheumatologists’**<br>**characteristics** | **Total** |
| --- | --- |
|  | n = 43 |
| **Demographics** |  |
| **Age, yr** | 39.6 (7.2) |
| **Women, n (%)** | 10 (23.3 %) |
| **Job title, n (%)** |  |
| Lecturer or lower | 36 (83.7 %) |
| Associate professor or higher | 7 (16.3 %) |

Note. Continuous variables are presented as medians and interquartile ranges (in square brackets or parentheses). Categorical variables are presented as frequencies and proportions (in parentheses). SLEDAI-2K: Systemic Lupus Erythematosus Disease Activity Index.

Fifty-one physicians responded to the questionnaire. Seven physicians were excluded because they were not listed as attending rheumatologists for SLE patients. The mean age was 42.4 (SD, 5.6) and 12 of them (23.5%) were women.

### Physician’s personality and comparison with the TIPI-J representative values

Among the attending physicians’ Big Five personality traits, agreeableness scores were the highest and conscientiousness scores were the lowest (Table 2). These characteristics are similar to those of representative values described in the TIPI-J development study. Compared to the representative values, attending physicians’ conscientiousness was significantly higher and their emotional stability was significantly lower.

**Table 2.** Rheumatologists’ personality scores and comparison with their representative scores.

| *Personality domain* | Rheumatologists' personality (n = 43) |  |  | The TIPI-J representative scores (n = 902) | *p-value\** |
| --- | --- | --- | --- | --- | --- |
|  | *Range* | *Median* | *Mean (SD)* | *Mean (SD)* |  |
| *Extraversion* | 3 - 14 | 8 | 8.09 (2.83) | 7.83 (2.97) | 0.55 |
| *Agreeableness* | 4 - 12 | 10 | 9.6 (1.88) | 9.48 (2.16) | 0.67 |
| *Conscientiousness* | 3 - 14 | 6 | 6.98 (2.47) | 6.14 (2.41) | 0.03 |
| *Emotional stability* | 3 - 13 | 7 | 7.77 (2.49) | 9.21 (2.48) | 0.0005 |
| *Openness* | 3 - 14 | 8 | 7.95 (2.45) | 8.03 (2.48) | 0.84 |

\*Unpaired t-test
Note. Representative scores were cited from the TIPI-J Development Paper. Scores were obtained from healthy university students from the four prefectures. [16]
TIPI-J: Japanese version of the Ten-Item Personality Inventory scale

### The relationship between the physician’s Big Five personality and trust in physicians

The relationship between physicians’ personality traits and patients’ trust in their rheumatologists is shown in Table 3. Trust in one’s rheumatologist increased with more ‘Extraversion’ and ‘Agreeableness’ personality (per 1-pt increase: 1.85 [95% CI 0.52, 3.19] and 2.24 [95% CI 0.86, 3.61], respectively). In contrast, ‘Conscientiousness’ personality was associated with reduced trust in one’s rheumatologist (per 1-pt decrease: 1.07 [95% CI −1.64, −0.49]). Disease duration > 20 years was associated with lower physician trust (−5.74 [95% CI −10.63, −0.86]).

**Table 3.** The associations of trust in patients’ attending rheumatologists with the rheumatologists’ Big Five personality traits and covariates.

|  | Corresponding<br>standardised ES | mean difference, point<br>estimate (95% CI) | p-value |
| --- | --- | --- | --- |
| **<u>Rheumatologists' characteristics</u>** |  |  |  |
| **Extraversion, per 1pt** | 0.12 | **1.85 (0.52 to 3.19)** | **0.008** |
| **Agreeableness, per 1pt** | 0.14 | **2.24 (0.86 to 3.61)** | **0.002** |
| **Conscientiousness, per 1pt** | -0.07 | **-1.07 (-1.64 to -0.49)** | **0.001** |
| Emotional stability, per 1pt | 0.0002 | 0.003 (-1.16 to 1.16) | 0.995 |
| Openness, per 1pt | -0.07 | -1.12 (-2.49 to 0.26) | 0.108 |
| Age, per 10 yrs | -0.02 | -2.54 (-7.67 to 2.59) | 0.323 |
| Women vs. Men | -0.05 | -0.77 (-6.38 to 4.83) | 0.781 |
| Job title (Assoc. Prof. or higher<br>vs. Lecturer or lower) | 0.08 | 1.33 (-5.83 to 8.48) | 0.71 |
| **<u>Patients' characteristics</u>** |  |  |  |
| Age, per 10 yrs | 0.004 | 0.66 (-0.63 to 1.95) | 0.307 |
| Women vs. Men | -0.14 | -2.27 (-10.15 to 5.62) | 0.564 |
| **Disease duration** |  |  |  |
| ≤5 yr |  | Reference |  |
| >5 yr to ≤10 yr | 0.18 | 2.87 (-0.96 to 6.7) | 0.138 |
| >10 yr to ≤20 yr | -0.05 | -0.79 (-4.51 to 2.94) | 0.671 |
| **>20 yr** | -0.36 | **-5.74 (-10.63 to -0.86)** | **0.022** |
| SLEDAI-2K, per 1pt | 0.01 | 0.16 (-0.26 to 0.57) | 0.449 |
| **Duration with their rheumatologist** |  |  |  |
| <6 m |  | Reference |  |
| ≥6 m to <1 yr | -0.005 | -0.08 (-8.71 to 8.56) | 0.985 |
| ≥1 yr to <3 yr | 0.2 | 3.24 (-2.93 to 9.42) | 0.295 |
| ≥3 yr | 0.19 | 2.98 (-2.9 to 8.85) | 0.312 |

A general linear model with cluster-robust standard estimation was fit with the inclusion of all variables listed above, with 43 patients' attending rheumatologists treated as a cluster. To calculate the corresponding standardised ES (Cohen's *d*), the point estimate was divided by the SD of the trust in the rheumatologist score.
Bold font indicates significance at $P < 0.05$ . ES: effect size; SLEDAI-2K: SLEDAI 2000

## DISCUSSION

This study examined the association between the Big Five personality traits of attending physicians and physician trust among patients with SLE. Our results showed that higher levels of extraversion and agreeableness of the attending physician were associated with higher levels of patients’ trust in their attending physicians; conversely, higher levels of conscientiousness of the attending physicians were associated with lower levels of patients’ trust in their attending physicians.

Several studies have investigated the impact of physicians’ Big Five personality traits on physicians’ medical practice, communication style, and patient ratings. Many of these studies reported positive effects for extraversion, openness, and agreeableness, whereas there were conflicting effects for conscientiousness. Physicians with higher conscientiousness and extraversion were more likely to choose intensive treatment for older leukaemia patients, [25] and pathologists with higher conscientiousness were more accurate in diagnosing lung cancer [26] During investigation of the psychosocial and lifestyle conditions of patients with depressive symptoms, physicians with higher openness were more likely to explore their medical history; however, physicians with higher conscientiousness were less likely to elicit patients’ wishes during the treatment plan phase [27]. Although the present study is the first to investigate the impact of rheumatologists’ Big Five personality traits on SLE patients’ trust in their attending physicians, it is noteworthy that the positive impact of agreeableness and the negative impact of conscientiousness were similar to the results of a previous study examining their associations with patient satisfaction. [14]

There are several possible explanations for how rheumatologists’ Big Five personality traits influence SLE patients’ trust in their attending physicians through their communication style. The reason underlying the high level of trust associated with highly agreeable rheumatologists is straightforward. Agreeableness is associated with the tendency to maintain pleasant and harmonious interpersonal relationships; [13] highly agreeable rheumatologists interact with their patients with compassion and respect. One study suggested that compassionate and patient-centred behaviour may help build trust in patients with SLE and rheumatoid arthritis. [28] There are several explanations for the higher level of patients’ trust in physicians with higher extraversion of physicians. One possible explanation is that physicians with extraversion are more sociable, likely to initiate conversations, and positively influence their patients. [13] In contrast, a lack of extraversion tends to keep physicians’ thoughts and feelings toward themselves, and the atmosphere around such a physician prevents patients from expressing what they need to say and perhaps impedes the formation of trusting relationships.[13] Medical students who were extremely shy, did not engage in non-verbal communication such as eye contact, and did not actively participate in conversations received poor ratings in their clinical performance, confirming that the poor rapport building was associated with a lack of extraversion. [29] While the reason for the lower trust in attending rheumatologists with higher conscientiousness, generally considered a good trait, is unclear, several hypotheses and observations support the present findings. Since conscientiousness is characterised by industriousness, such as the thorough completion of tasks, self-control to delay short-term gratification, and responsibility to fulfil obligations and follow rules, [30] conscientiousness predicted higher performance in medical skills, such as Objective Structured Clinical Examinations (OSCEs) and surgery. [31] However, conscientiousness is observed to have a potential disadvantage, [31] and overly conscientious physicians may adhere to norms and adversely influence patient interaction. An SLE patient’s report by their rheumatologist lacked the symptoms presented by the patient because the rheumatologist was too engrossed with typical symptoms from medical text or experiences gained with other patients. [32] Such a patient’s perception of difficulty in communicating with their rheumatologist may cause the patient to feel that the rheumatologist does not believe their reports, [2] which can result in a fragile physician-patient relationship.

The clinical implications of this study are noteworthy. First, the integration of communication education, specifically addressing the doctor-patient relationship, into medical school or specialized training programs may modify the impact of a physician’s personality on rapport with their patients. Although personality traits are often considered invariant, previous studies have shown that while they are heritable and remain stable into adulthood,[33] they can be modified by life events and through intentional efforts.[34] Second, recognising and reflecting on one’s personality traits can provide opportunities to apply the traits in communicating with patients. For example, a high conscientiousness can provide an opportunity for the physician’s self-reflection on whether they are overly adhering to treatment guidelines without the interference of patient’s preferences. A previous study suggested that a personality trait persisting in reducing glucocorticoids to achieve low disease activity in SLE could make it difficult to achieve that aim.[35] Furthermore, given that agreeable personality was slightly protective against medical negligence claims[36], a retrospective analysis of a physician’s personality traits in doctor-patient disputes might provide an opportunity to attribute the cause of the dispute to a specific communication style and find a way to resolve the dispute.

This study had several strengths that are noteworthy. First, this was a multi-centre study; therefore, the observed impact of rheumatologists’ personality traits on trust is generalisable to similar academic medical centres, instead of reflecting the specific culture of a particular institutional department. Second, we adjusted for the physician-level clustering of trust with an appropriate multilevel analysis, based on the merging of the rheumatologist-by-rheumatologist basis of data from the attending rheumatologists and those from a large enough number of patients. As a result, the observed association between rheumatologists’ personality traits and trust was not reflective of a particular rheumatologist’s traits.

This study had some limitations. First, the Big Five personality traits were rated via a self-report and, therefore, not objective. However, the consistency between self-reported and objective assessments was verified in a previous study. [37] Second, unlike other scales of trust in one’s physician, [7] the scale used in this study does not include confidentiality. Consequently, it may not be able to measure the aspect of trust in conscientious physicians that is maintained as a result of adherence to the principles of confidentiality.

Our finding that SLE patients’ trust in their physicians is related to their physicians’ Big Five personality trait components indicates that such personality traits potentially influence physicians’ dialogue patterns and underlie the formation of the doctor-patient relationship. It is necessary to incorporate behaviours and dialogue that are effective in intentionally maintaining the therapeutic relationship, as the attending physician reflects on whether conscientiousness is excessive or extraversion or agreeableness is insufficient.

## Data Availability

The datasets generated and/or analysed during the study are available from the corresponding author upon reasonable request.

## ACKNOWLEDGMENTS

We would especially like to thank Hiroko Nagasato, Kumi Sasaki, Yukari Hosaka (Showa University), and Miyuki Sato (Fukushima Medical University) for their support. A part of this study was presented at the 67th Annual General Assembly and Scientific Meeting of the Japan College of Rheumatology.

