## Supplemental Table1 for "The impact of attending rheumatologist’s Big Five personality traits on systemic lupus erythematosus patients’ trust in their rheumatologist: the TRUMP^2^-SLE project"

**Supplementary Table 1. Ten-Item Personality Inventory Scale (TIPI-J)**

| Instruction sentence | Here are a number of personality traits that may or may not apply to you. Please write a number next to each statement to indicate the extent to which you agree or disagree with that statement. You should rate the extent to which the pair of traits applies to you, even if one characteristic applies more strongly than the other . |
| --- | --- |
|  | I see myself as |
| Question 1 | Extraverted, enthusiastic. |
| Question 2 | Critical, quarrelsome. |
| Question 3 | Dependable, self-disciplined. |
| Question 4 | Anxious, easily upset. |
| Question 5 | Open to new experiences, complex. |
| Question 6 | Reserved, quiet. |
| Question 7 | Sympathetic, warm. |
| Question 8 | Disorganized, careless. |
| Question 9 | Calm, emotionally stable. |
| Question 10 | Conventional, uncreative. |
| Response options for questions | (Disagree strongly / Disagree moderately / Disagree a little / Neither agree nor disagree/ Agree a little/ Agree moderately/ Agree strongly) |
